# Trends in advanced HIV disease, treatment interruption, and viraemia in KwaZulu-Natal, South Africa

**DOI:** 10.1101/2025.11.06.25339741

**Authors:** Sanele S. Mbeje, Lilishia Gounder, Johan S. van der Molen, Lungile Hobe, Thulani Ngwenya, Mlungisi Khanyile, Kwena Tlhaku, Benjamin Chimukangara, Thokozani Khubone, Yukteshwar Sookrajh, Sharana Mahomed, Jennifer A. Brown, Nigel Garrett, Jienchi Dorward, Lara Lewis

## Abstract

Advanced HIV disease (AHD), treatment interruption and viraemia remain key challenges to effectiveness of HIV programmes in South Africa. Understanding the trends of these measures and their variation across geographic regions can inform more targeted and responsive interventions.

We conducted a retrospective cohort study using routine, de-identified data from TIER.Net, a national HIV electronic register. We curated data from 116 primary care clinics in eThekwini, uMgungudlovu, and uMkhanyakude Districts in KwaZulu-Natal province. We included people living with HIV aged ≥16 years, who initiated or collected ART between 1 July 2018 and 30 June 2023. We examined trends in annual proportion of AHD, treatment interruption and viraemia, and mapped the clinic-level proportions of these outcomes using inverse distance weighted interpolation maps.

Among 123,473 clients initiating ART with CD4 count measurements, the overall prevalence of AHD remained stable over the period (20.0%-22.2%). There was an overall decline in treatment interruptions from 13.2% to 11.3% among the 544,066 clients who had scheduled visits. Of the 446,899 clients who had viral load results, overall viraemia ≥50 copies/mL increased from 12.9% to 20.2%, whereas viraemia ≥1000 copies/mL remained stable at approximately 6%. Spatial analyses highlighted geographic disparities in AHD, treatment interruption, and viraemia across and within the districts.

While declines in treatment interruptions are promising, the persistence of AHD and viraemia (≥50 copies/mL) require further investigation and highlight ongoing challenges for HIV control. The observed spatial variation across districts underscores the need of geographically tailored interventions to strengthen programmatic effectiveness, particularly in hyper-endemic settings like KwaZulu-Natal.

## Introduction

Human immunodeficiency virus (HIV) continues to be one of the most challenging public health issues in South Africa and globally. Despite significant advances, SA remains the epicentre of the HIV epidemic[1, 2]. The country hosts one of the largest HIV treatment programmes and has made progress in identifying untreated people living with HIV (PLWH), expanding access to testing and care services, and improving viral suppression[1]. However, HIV prevalence is not evenly distributed across regions. The KwaZulu-Natal province has one of the highest HIV prevalences in the country, estimated at 16% in 2022, with some of the districts in this province reporting prevalence rates as high as 20%[3].

Early initiation of antiretroviral therapy (ART) has reduced morbidity, mortality and HIV transmission[4, 5]. In 2015, the World Health Organization (WHO) recommended ART for all PLWH, regardless of the WHO clinical stage or CD4 count, which led to the Universal Test and Treat (UTT) policy being implemented in SA in 2016[6]. This guidance has contributed to stabilizing HIV incidence, improving survival rates and reducing mortality[1]. Despite these gains a substantial number of individuals in SA continue to present with advanced HIV disease (AHD), defined as having a CD4 cell count below 200 cells/mm^3^ or WHO clinical stage III/IV at ART initiation, indicative of late initiation[7-9]. The 2017 WHO guidelines advised that clients with AHD should receive a comprehensive care package that includes screening, co-infection prophylaxis treatment, rapid ART initiation, and intensified adherence support[9]. Baseline CD4 count is key to identifying clients at risk of opportunistic infections, and for understanding the clinical care needs for those who present late[10]. While the national data shows a decline in the prevalence of AHD over time, recent findings indicate that approximately 20% of clients present with AHD[2, 11]. Moreover, substantial regional disparities persist, with provincial rates ranging between 11.5% and 26.6% in recent years[12, 13], underscoring the uneven burden.

Since the introduction of UTT, the number of clients eligible for ART has increased. Adherence to ART reduces onward transmission risk of those living with HIV as well as the probability of CD4 count decline[14].

Therefore, ensuring that PLWH are enrolled and retained in care, and adherent to ART is crucial for better HIV outcomes[15]. Despite this evidence, treatment interruption remains a major challenge, contributing to morbidity, mortality, and transmission[15, 16]. Studies conducted in various regions of SA between 2013 and 2022 have observed disengagement from care between 14.7% to 38.8% of clients within an up to two-year period[17-19]. These findings highlight that disengagement in care continues to be an important challenge despite expanded access to treatment.

Viral load (VL) monitoring is a key component of ART success, as achieving an undetectable VL is associated with long-term health benefits and eliminates the risk of onward sexual transmission[14]. However, viraemia (often due to poor adherence, or disengagement from care) continues to pose a challenge. South African National Health Laboratory Service (NHLS) data from 2013 to 2022 showed a rise in low-level viraemia VL (50–999 copies/mL), from 15.3% in 2018 to 20.5% in 2022, while high level viraemia (≥1000 copies/mL) declined during the same period[20]. In Mpumalanga province, 2019 data indicated that 13.8% of women had low-level viraemia (50–999 copies/mL), while 14.7% had VLs ≥1000 copies/mL[21].

In this study we focused on three persistent challenges that continue to undermine effectiveness of HIV programmes, namely AHD, treatment interruption, and viraemia, aiming to analyse their temporal trends and spatial patterns in three high-burden districts of KwaZulu-Natal, South Africa. The prevalence of these measures varies over time as well as across provinces and sub-districts, making spatial and temporal monitoring critical for informing more focused and effective interventions to improve HIV outcomes.

## Methods

### Study design, setting and population

We conducted a retrospective cohort study including data from 116 primary healthcare clinics managed by eThekwini Municipality, uMgungudlovu District, and Mseleni/Bethesda Hospital in uMkhanyakude District in KwaZulu-Natal province. We included all PLWH aged 16 years and older initiating and collecting ART at participating clinics between 01 July 2018 and 30 June 2023.

HIV care service delivery in South Africa includes the provision of free ART in public healthcare facilities along with a wide range of clinical assessments and laboratory evaluations[22], including VL and CD4 count testing[22, 23]. As per 2019 guidelines, VL testing was done at 6 and 12 months after ART initiation and annually thereafter for those who remained virally suppressed, while CD4 count testing was done at initiation, at 12 months on ART and when clinically indicated[22].

### Data source and data management

We analysed routinely collected de-identified electronic health record data from Tier.Net, an electronic register used to maintain tuberculosis and HIV client information in public healthcare facilities in South Africa. It contains clients’ demographics, records of ART use, clinic visits, laboratory tests (CD4 counts and VLs), tuberculosis treatment history, and clinical outcomes[24, 25].

### Outcomes

Study outcomes were the proportion of clients with AHD, treatment interruption, and viraemia, and were measured annually. We estimated AHD proportions using data from all clients initiating ART who had CD4 count data measured within 180 days before to 30 days post ART initiation. Clinical staging data was not available in our dataset, only CD4 count < 200 cells/mm^3^ was used to define AHD. For clients with multiple CD4 counts within the defined window, we prioritized pre-initiation measurements closest to the initiation date, and then post-initiation measurements.

To evaluate treatment interruption, we included all PLWH on ART who had at least one visit scheduled during the study period. We defined treatment interruption as missing any scheduled visit by more than 90 days, irrespective of duration on ART and any previous interruptions. We calculated the proportion of clients who experienced at least one treatment interruption in a year. Individuals who were transferred to another clinic or died within 90 days of a scheduled visit were not defined as having a treatment interruption.

For measurement of viraemia, we included all PLWH who had been on ART for more than 5 months. Among those who had at least one VL result in a year, we calculated the proportion of clients who had VL ≥50 copies/mL and VL ≥1000 copies/mL in a year. If a client had multiple VL tests in a year, the highest VL was selected.

### Statistical Analysis

We used descriptive statistics to summarize clinic-level characteristics in the most recent reporting period (01 July 2022 – 30 June 2023) using medians and interquartile ranges (IQRs), or counts and percentages. Clinic-level characteristics such as median age, sex distribution were generated by aggregating client-level data annually and per clinic. We estimated clinic size as the number of clients who had at least one visit at that clinic within the study period and assessed socioeconomic context of the clinics using ward-level data. We used the 2011 ward-level South African multidimensional poverty index (SAMPI) from Statistics South Africa to assess area-level deprivation, the most recent census-based metric available at the time of analysis[26]. SAMPI scores range from 0 to 1, where higher scores indicate greater level of deprivation[26]. We categorized SAMPI scores into quintiles based on cut-off values derived from the KwaZulu-Natal ward-level SAMPI distribution, to classify whether clinics are located in areas ranging from the least deprived (Q1) to the most deprived (Q5). Each participating clinic was linked to a ward using its global position system (GPS) coordinates.

We used line plots and bar charts to visualize trends of our study outcomes. To estimate the clinic-level relative risk (RR) with 95% confidence intervals (CIs) of AHD, treatment interruption and viraemia ≥50 copies/mL, we employed a log-binomial generalized linear mixed model (GLMM) with random slope and clinic-specific random effects on the intercept to account for the heterogeneity in outcomes by clinic, and correlated observations within clinics[27-29]. Clinic-level characteristics of median age, clinic size quintiles, SAMPI quintiles, male proportion and year (of ART initiation, scheduled visit or VL result) were included as fixed effects. Client-level characteristics were summarized at the time of ART initiation, time of VL testing and for treatment interruption at first visit within each year. All analyses were performed using R software (version 4.4.0).

### Geospatial mapping

To visualize spatial patterns of each outcome across the five-year period, we calculated crude clinic proportions and generated geospatial maps using QGIS software version 3.40.0[30]. Mapping was based on a vector layer of clinic records containing GPS coordinates (S1 Fig), clinic names, district, ward identifier and proportions of each outcome, and the shapefiles delineating 2020 local municipality and ward boundaries in KwaZulu-Natal sourced from the Municipality Demarcation Board website[31].

We generated inverse distance weighting (IDW) interpolation maps to visualize the proportions of each outcome per district. IDW is a spatial interpolation technique that estimates values at unsampled locations (e.g. areas surrounding the participating clinics) based on known values, assigning weights proportional to the distance between clinics. It operates on the principle that spatial phenomena at proximate locations are more similar than those further apart[32-35].

### Ethical approval

This study was approved by the University of KwaZulu-Natal Biomedical Research Ethics Committee (BE646/17), eThekwini Municipality Research Committee, and the KwaZulu-Natal Department of Health’s Provincial Health Research Ethics Committee (KZ_201807_021), with a waiver for informed consent for analysis of anonymized routinely collected data.

## Results

Among 116 clinics included in analysis, the median clinic size between July 2022 and June 2023 was 4,212 (IQR 2,677–6,105), Table 1. The median age was 39 (IQR 38, 40) years and of males was 31.7% (IQR 30.3%, 33.6%). Approximately one in every five clinics (23, 19.8%) were located in the most deprived quintiles (Q4 and Q5). Geographically, half of the clinics were located in eThekwini with 38.8% in uMgungundlovu, and 11.2% in uMkhanyakude.

**Table 1.** Characteristics of clinics with HIV data analysed between July 2022 and June 2023.

| Characteristic | N = 116,<br>Median [IQR]; |
| --- | --- |
| Clinic size | 4,212 [2,677; 6,105] |
| Median age | 39.00 [38.00, 40.00] |
| Proportion of male | 0.317 [0.303, 0.336] |
| Multidimensional Poverty Index quintiles | n (%) |
| Q1 (least deprived) | 46 (39.7%) |
| Q2 | 32 (27.6%) |
| Q3 | 15 (12.9%) |
| Q4 | 13 (11.2%) |
| Q5 (most deprived) | 10 (8.6%) |
| District | n (%) |
| eThekweni | 58 (50.0%) |
| uMgungundlovu | 45 (38.8%) |
| uMkhanyakude | 13 (11.2%) |

### Advanced HIV disease

We identified 199,969 clients who initiated ART, of whom 164,752 were newly initiated and were included in the AHD analysis. Of these, 41,279 did not have CD4 count data (Fig 1A). While the overall number of ART initiations decreased from 45,780 (July2018–June2019) to 22,347 (July2022–June2023), the percentage of clients with CD4 count results recorded at ART initiation improved from 71.6% to 78.0% over the period (Fig 2). The overall prevalence of AHD remained unchanged, ranging between 20.0% and 22.2%. Similar stable trend of AHD was observed at a district level (S2 Fig), although fluctuating trends were observed at a sub-district level (S3 Fig).

**Fig 1.**
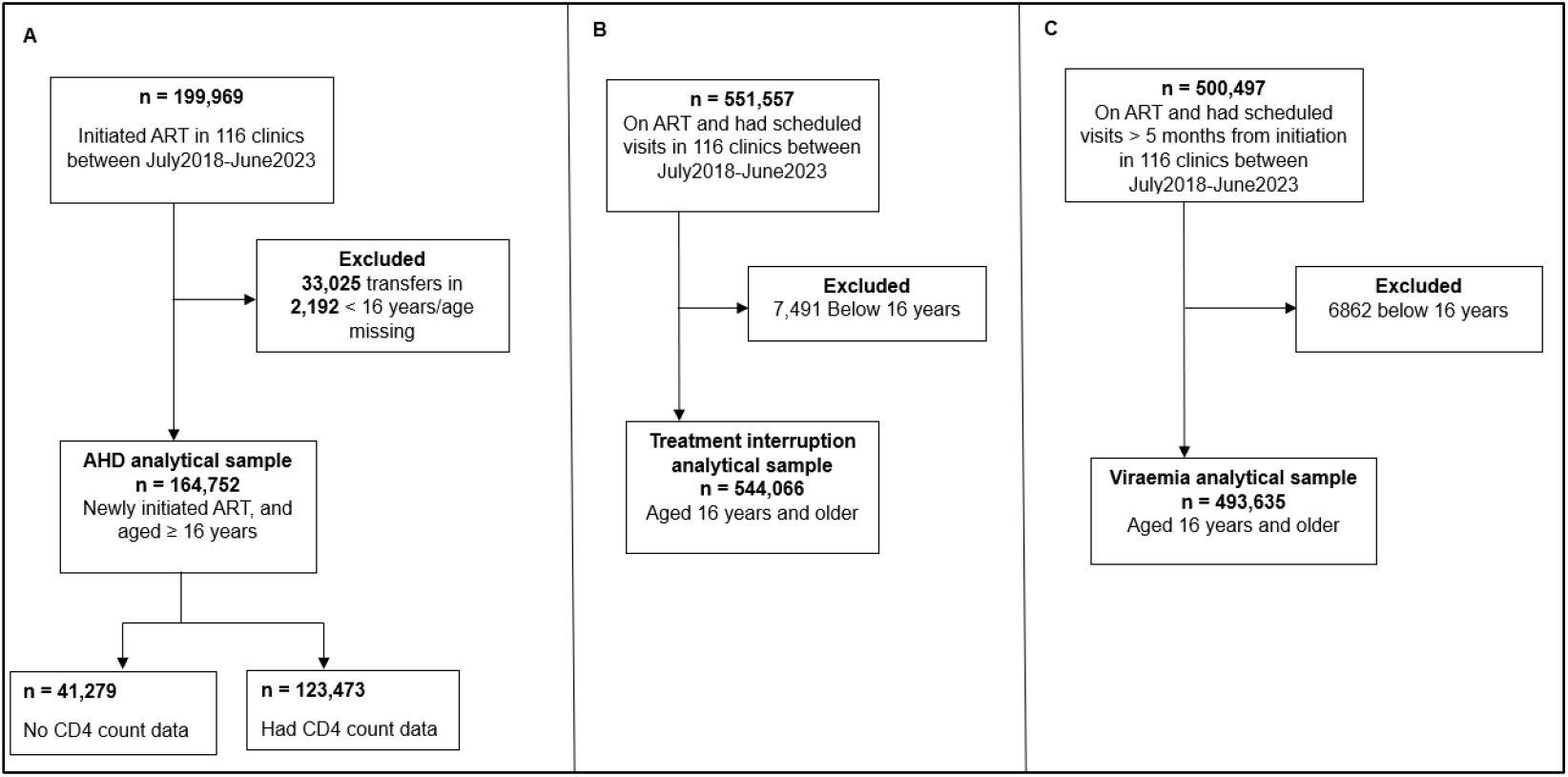
Flowchart for each study outcome. A: advanced HIV disease, B: treatment interruption, and C: viraemia.

**Fig 2.**
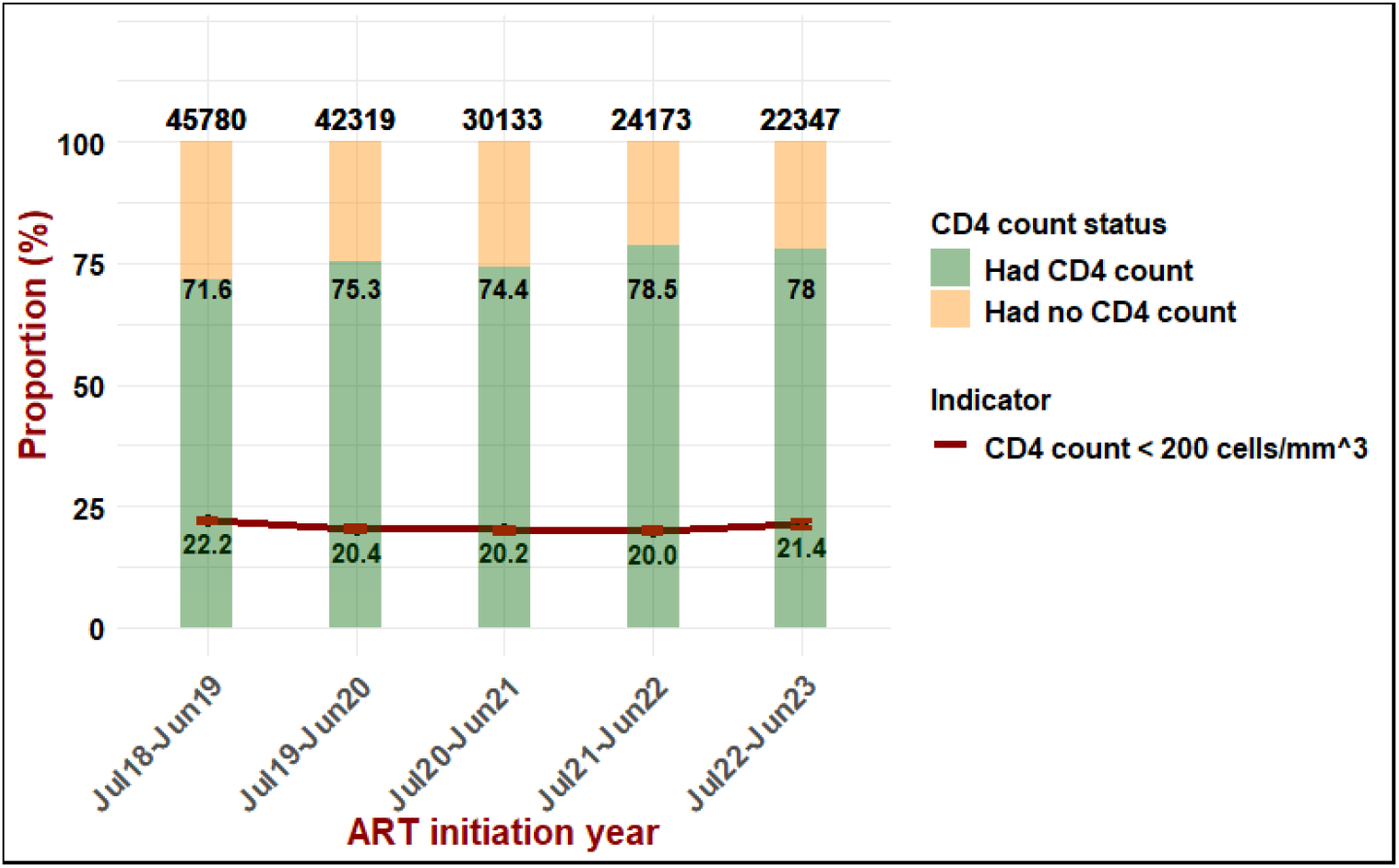
Overall number of ART initiators and prevalence of AHD among clients who had CD4 count across three districts.

The clinic-level prevalence of AHD across the five-year period ranged from 15% to 32% (Fig 3) and varied within each district. In uMgungundlovu (Fig 3a), most clinics recorded a prevalence of 20% or higher, with notably higher levels in the western region. In eThekwini (Fig 3b), the highest AHD prevalence was observed in the southern and western regions, while in uMkhanyakude (Fig 3c), the prevalence was higher in the northern, western and far southern regions.

**Fig 3.**
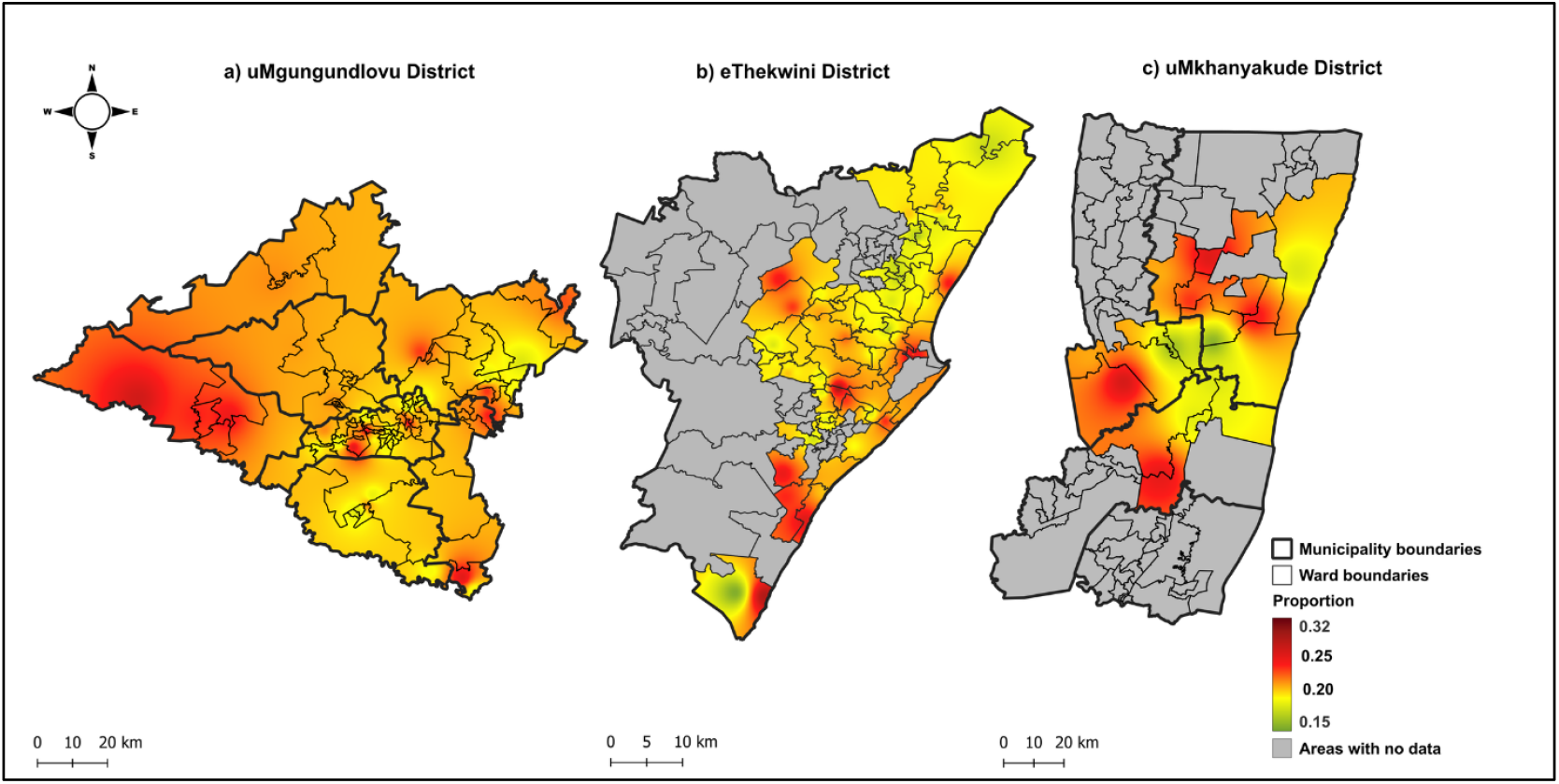
Spatial distribution in prevalence of AHD in three districts of KwaZulu-Natal.

The risk of AHD declined slightly over the years (RR:0.98, 95%CI: 0.97-0.99), Table 2. Clinics serving older populations (RR: 1.07, 95%CI: 1.04-1.09) and an increase in proportions of males (RR: 1.01, 95% CI: 1.00-1.01) was associated with increased risk of AHD.

**Table 2.** Adjusted relative risk of clinic-level prevalence of AHD, treatment interruption, viraemia.

| Characteristic | AHD |  | Treatment interruption |  | Viraemia |  |
| --- | --- | --- | --- | --- | --- | --- |
|  | RR (95% CI) | P-value | RR (95% CI) | P-value | RR (95% CI) | P-value |
| Median age (standardized) | 1.07 (1.04,1.09) | <0.001 | 0.98 (0.96,1.00) | 0.101 | 0.92 (0.91,0.94) | <0.001 |
| Proportion of male (per 1% change) | 1.01 (1.00,1.01) | 0.001 | 0.94 (0.93,0.95) | <0.001 | 1.02 (1.01,1.02) | <0.001 |
| MPI (quintiles) | 1.00 (0.98,1.02) | 0.981 | 0.95 (0.91,1.00) | 0.031 | 1.01 (0.98,1.04) | 0.610 |
| Clinic size (quintiles) | 0.98 (0.97,1.00) | 0.074 | 0.99 (0.95,1.03) | 0.585 | 0.96(0.94,0.99) | 0.004 |
| Year | 0.98 (0.97,0.99) | 0.001 | 0.99 (0.96,1.01) | 0.270 | 1.12 (1.09,1.14) | <0.001 |

RR = Relative Risk, CI = Confidence Interval, MPI = Multidimensional Poverty Index, AHD=advanced HIV disease

### Treatment interruptions

Among 551,557 clients who were on ART and had at least one scheduled visit during the study period, 544,066 were 16 years or older and were included in the analysis (Fig 1B). The number of clients with scheduled visits increased from 343,796 in July2018–June2019 to 385,306 in July2022–June2023 (Fig 4). The proportion of treatment interruption declined from 13.2% in July2019–June2020 to 11.3% by July2022– June2023. Slightly varied trends were observed at a district level (with eThekwini declining from 13.8% to 10.8%) and at a sub-district level (S4 and S5 Figs).

**Fig 4.**
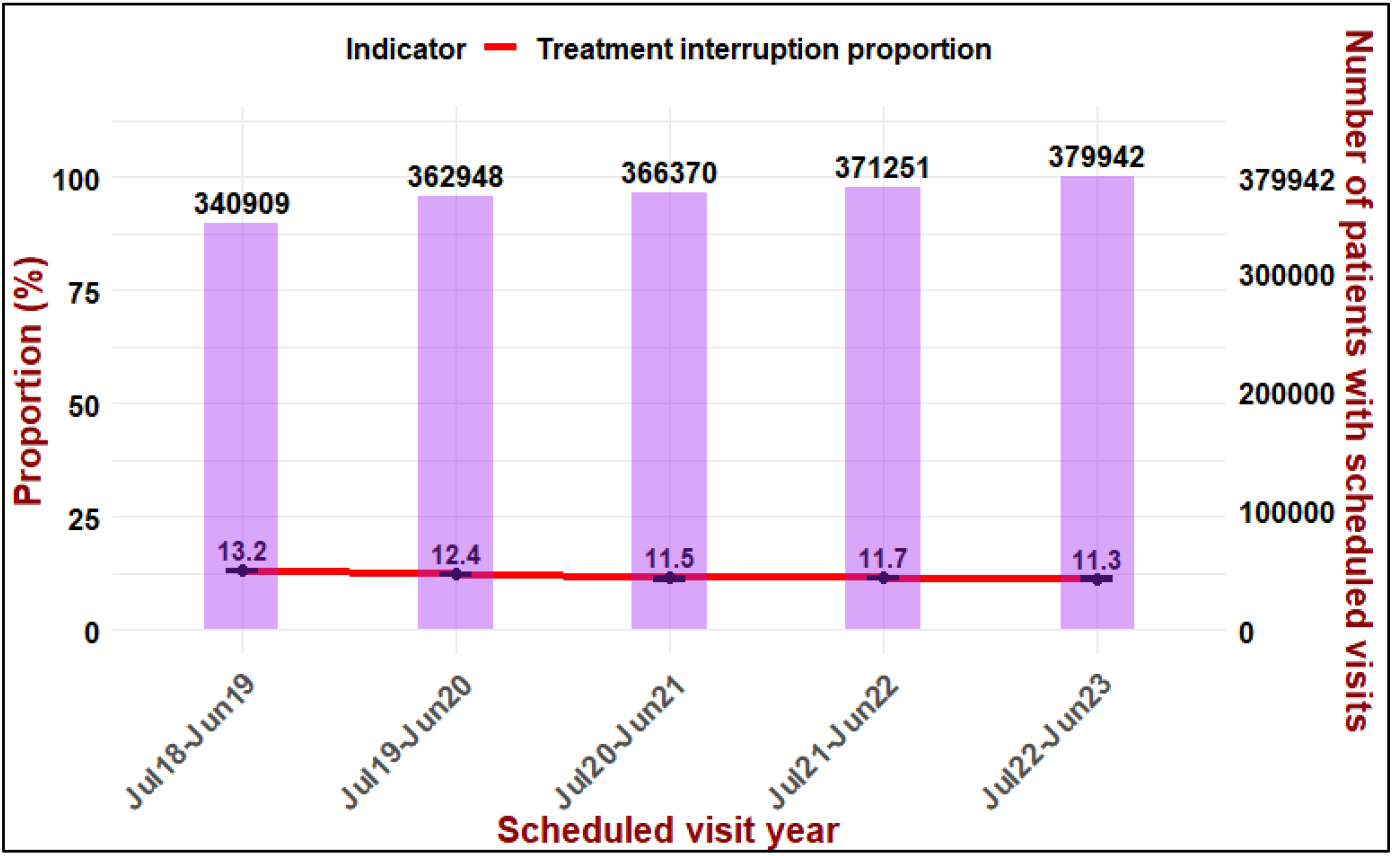
Overall number of PLWH who had scheduled visits and proportion of treatment interruption across three districts.

The proportion of treatment interruption varied across the three districts (Fig 5). In uMgungundlovu, it was generally consistent across most clinics, with higher levels of between (15% and 30%) in parts of the western and southern regions. In eThekwini it was highest in clinics located in the central region (between 15% and 20%). Treatment interruption proportions varied across clinics in the uMkhanyakude. Those located in the western and far southern region had the highest levels, ranging from 15% to 25%.

**Fig 5.**
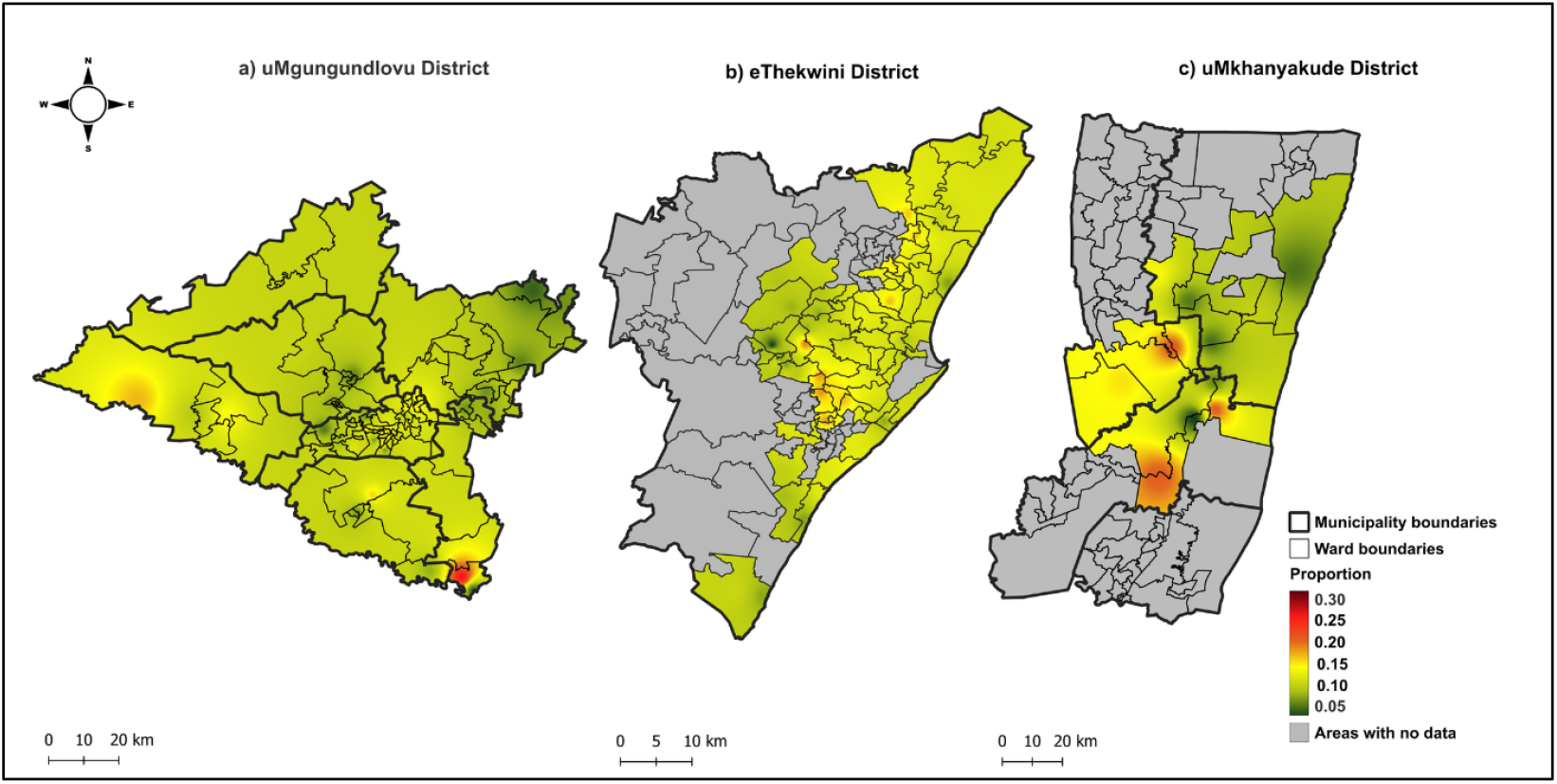
Spatial distribution in the proportion of treatment interruption in three districts of KwaZulu-Natal.

In the adjusted GLMM (Table 2), an increase in proportion of males lowers the risk of treatment interruption (RR: 0.94, 95%CI: 0.93-0.95), and the risk was reduced for clinics located in the least deprived areas (RR: 0.95, 95%CI: 0.91-1.00).

### Viraemia

We identified 500,497 clients with at least one scheduled visit more than 5 months from initiation, and 493,635 clients (≥16 years old) were considered for the analysis (Fig 1C). The proportion of clients who had VL results was 76.9% in July2018–June2019 and increased to 82.7% in July2022–June2023 (Fig 6). While the proportion of clients who had high-level viraemia (≥1000 copies/mL) remained stable at approximately 6% across all years, the proportion of viraemia (≥50 copies/mL) increased from 12.9% to 20.2%. eThekwini district, experienced an upward trend in viraemia (≥50 copies/mL) while varying trends were observed in uMgungundlovu and uMkhanyakude Districts (S6 Fig).

**Fig 6.**
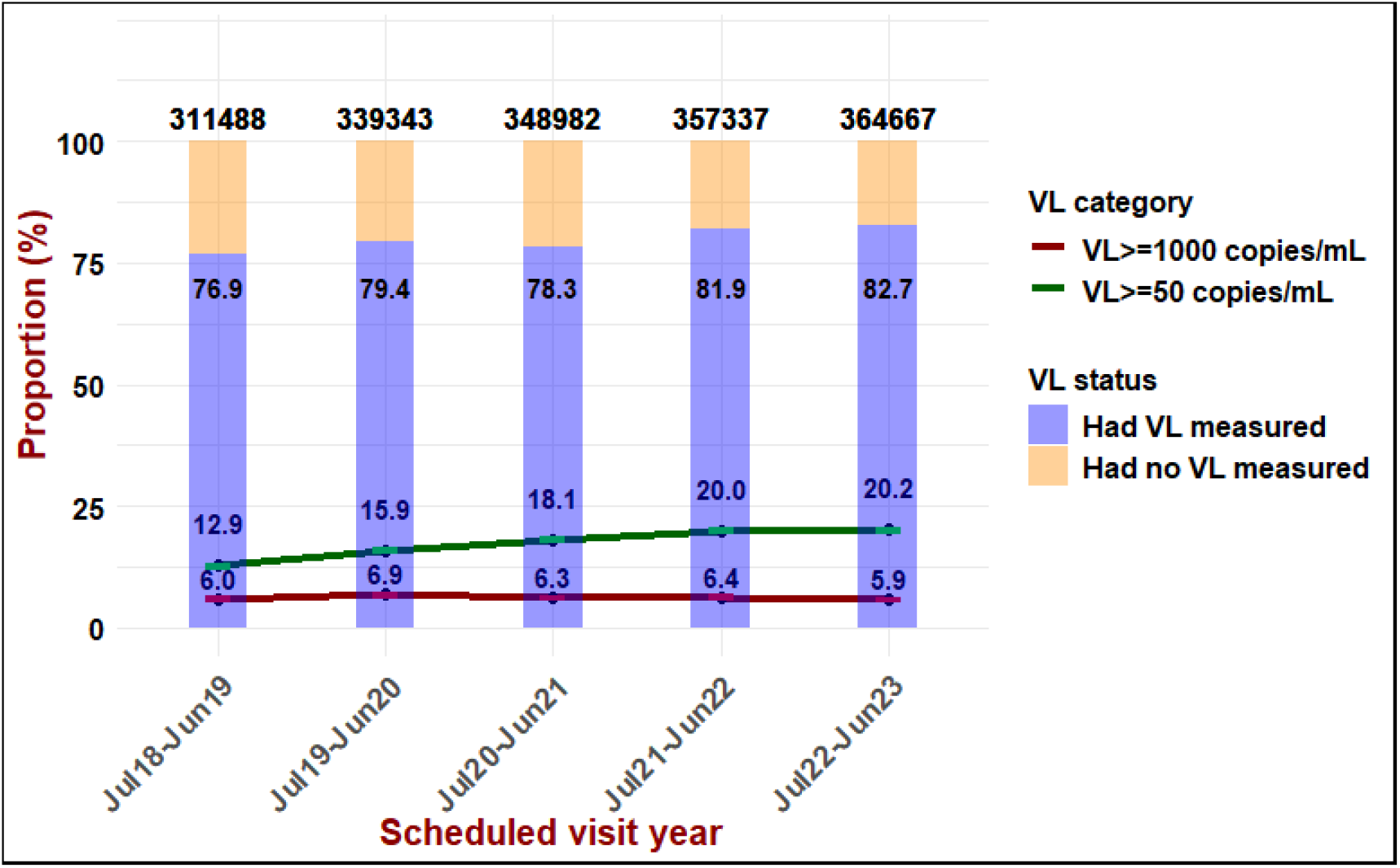
Overall number of PLWH who had scheduled visits more than 5 months from ART initiation and proportion of viraemia ≥50 copies/mL and ≥1000 copies/mL across three districts. VL, viral load; mL, millilitre.

High proportions of viraemia (≥50 copies/mL) were observed in clinics located in the central and north-eastern region of uMgungundlovu ranging from 15% to 30%, and in the northern and southern region of eThekwini between 15% and 25% (Fig 7). However, it was generally consistent across clinics in uMkhanyakude.

**Fig 7.**
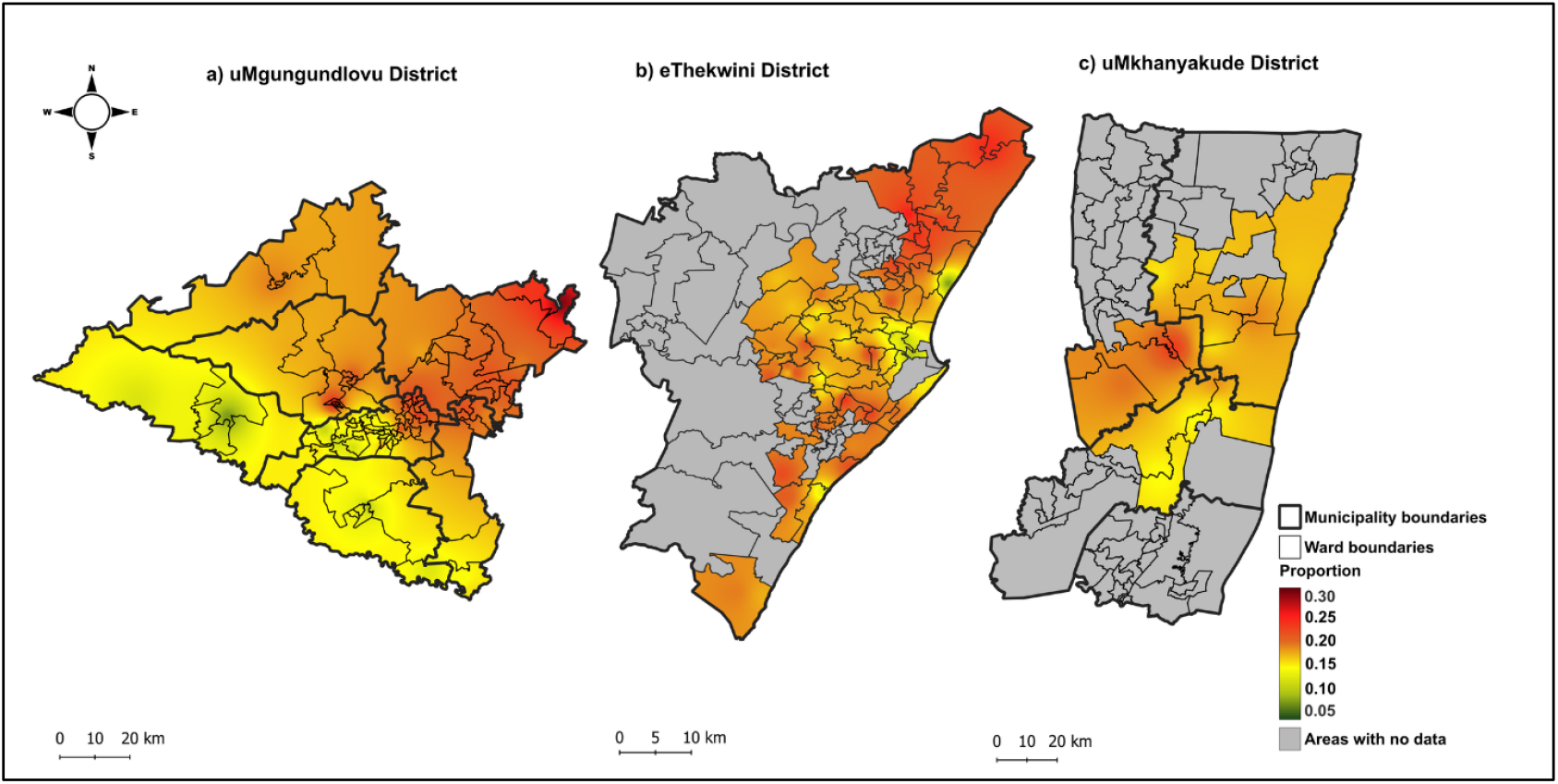
Spatial distribution in the proportion of viraemia (≥50 copies/mL) in three districts of KwaZulu-Natal.

At a clinic-level, the proportion of viraemia (≥50 copies/mL) increased significantly over the years (RR: 1.12, 95%CI: 1.09-1.14), Table 2. Clinics serving older clients had a reduced risk of viraemia (RR: 0.92, 95% CI: 0.91-0.94), while an increase in proportion of males was associated with an increased risk (RR:1.02 95%CI:1.01-1.02). However, an increase in the number of clients at a clinics was associated with a reduced risk (RR: 0.96, 95% CI: 0.94-0.99).

## Discussion

In this study, we examined temporal and spatial variation in the prevalence of AHD, treatment interruption, and viraemia, three key measures of HIV programmatic performance, using data from the KwaZulu-Natal province in South Africa between 2018 and 2023. We also explored whether temporal changes in prevalence are associated with clinic characteristics such as median clinic age, sex proportions, clinic size and clinic area wealth quintiles. The findings of this analysis can be used to inform more targeted interventions to improve HIV service delivery in KwaZulu-Natal and other hyper-endemic settings.

We found that AHD prevalence remained stable over the five-year period at approximately 20-22%, aligning with recent studies reporting AHD between 18.2% to 28.8% at a national and provincial level between 2017 and 2023[11, 36]. Spatial analysis revealed geographic disparities in AHD across and within districts, offering insights often masked by national or regional averages. In a clinic-level analysis, we found that an increase in proportion of males and aging of clients in clinics was associated with increased risk of AHD. This is consistent with other studies where older age and male sex were reported to be at high risk[2, 36]. Gender disparity is likely due to men’s lower engagement in HIV testing and treatment, driven by stigma and suboptimal health-seeking behaviors[37, 38]. Taken together, these results highlight that despite improved ART access, late presentation remains a challenge. To further reduce AHD among ART naïve clients, strategies must be more targeted given uneven distribution and focus on early HIV testing and linkage to care, especially among men[36].

Our definition of treatment interruption aligns with commonly used measures of disengagement from care[19, 39-42], however some studies have used longer thresholds than ours (e.g., >180 days late for a visit)[17, 43]. Studies have also assessed loss-to-follow-up at specified follow-up intervals since ART initiation (e.g., at 6, 12, 18, 24 months or more)[16, 39, 41, 42], whereas our analysis examined interruptions over time, and regardless of prior interruptions and time on ART. In the literature, disengagement from care varied between studies done in SA, ranging between 14.7% and 50.3% depending on the time on ART[17, 19, 41, 42], and period used for analysis. We observed a slight overall decline in treatment interruption, from 13.2% to 11.3% between 2018 and 2023. We found that treatment interruptions differ across and within districts. Interestingly, we observed that an increase in proportion of male clients in a clinic was associated with reduced risk of interruption. These findings differ from most existing literature reported in SA and elsewhere, where male sex tends to increase the risk of disengagement from care[15, 17, 19]. However, our analysis was conducted at a clinic-level, which may explain this discrepancy. A study conducted in SA between 2014 and 2018 showed that males initiating ART at male-focused clinics had reduced risk of dropping out of care[38], suggesting that the protective association we observe may be linked to unmeasured clinic-level factors. Importantly, this unexpected association could also reflect biases introduced by aggregating data, where group-level may not align with individual-level patterns[44]. Contrary to other studies which reported that older age is associated with reduced risk of disengagement from care[15], in this study age was not a significant factor.

Over the five-year period, we observed an overall increase in viraemia (≥50 copies/mL), rising from 12.9% to 20.2%. In contrast, viraemia (≥1000 copies/mL) remained stable at approximately 6%. A similar trend of increasing rates of low-level VL (50-999 copies/mL) both at regional and national level, alongside a decline in high-level viraemia over time has been reported by recent studies[20, 32]. These findings may reflect promising improvements in preventing sustained high-level viraemia, partly attributable to the roll-out of dolutegravir-based treatment in late 2019[22, 45], while highlighting a growing burden of low-level viraemia. Our findings also reveal varying trends at a district and sub-district level, with eThekwini showing an upward trend in viraemia (≥50 copies/mL). We also found that the distribution of viraemia varies across the districts. At a clinic-level, we showed that larger clinics serving younger clients were associated with reduced viraemia risk. However, an increase in the proportion of males tended to increase the risk of viraemia (≥50 copies/mL). While our findings differ from previous studies reporting that younger age is associated with an increased risk of viral rebound[46, 47], they align with other studies where male sex was associated with increased risk of viraemia[47, 48].

The size of our cohort (incorporating approximately half a million people on ART, drawn from both rural and urban public sector settings) and the length of the longitudinal data employed enhances the generalizability of our findings and is a key strength of the analysis. However, there are limitations to the use of TIER.Net data that must be highlighted. “Silent transfers” are not captured in TIER.Net and will have led to an over-estimation of treatment interruptions and misclassification of ART naïve clients[49]. VL and CD4 count data in TIER.Net is also not complete, although a recent study (not yet published) showed that the VL data quality has improved across all levels of VL (>50 copies/mL and >1000 copies/mL)[50]. Although CD4 count data remains under-captured, the degree of under-capture has not been associated with CD4 count level and therefore should not impact on the interpretation of our findings[50]. A final limitation of our analysis was that we did not have data on client residential area and so we used clinic coordinates to map the outcomes of the study. Residential location could have supported a more accurate estimation of clinic catchment areas as clients do not always visit clinics closest to their place of residence.

## Conclusion

In this study we demonstrated that treatment interruptions were fairly low, however AHD and viraemia (≥50 copies/mL) continue to pose a challenge to reducing HIV burden in SA. Our findings also revealed geographic heterogeneity in these measures. The persistent burden of AHD, increasing viraemia (≥50 copies/mL), and marked spatial variation across districts underscores the need for geographically tailored interventions to strengthen programmatic effectiveness in hyper-endemic settings like KwaZulu-Natal.

## Data Availability

The data used in this analysis cannot be publicly shared because of legal and ethical requirements regarding the use of routinely collected clinical data in South Africa.

## Abbreviations

(AHD): Advanced HIV disease
(ART): Antiretroviral therapy
(HIV): Human immunodeficiency virus
(PLWH): People living with HIV
(TIER.Net): Three Interlinked Electronic Register
(UTT): Universal test and treat
(VL): Viral load

## Author contributions

JD, NG, LL, and YS conceived the analysis. JD, NG, LL, SM, MK and KT were responsible for various components of the project administration. TK, YS, LH, TN, and MK oversaw data collection. JD, NG, TK, LL, and JSvdM oversaw data curation. SSM, JD, LL, JSvdM, and LG analysed the data with inputs from BC and JAB. SSM drafted the manuscript. All authors critically reviewed and edited the manuscript and consented to final publication.

## Acknowledgements

We would like to thank all staff and clients at eThekwini Municipality, uMgungudlovu District, Mseleni Hospital and Bethesda Hospital primary care clinics.

## Supporting information

**S1 Fig. Study area, sub-districts and clinic locations**.

**S2 Fig. Overall number of ART initiators and prevalence of AHD among clients who had CD4 count by district**.

**S3 Fig. Overall number of ART initiators and prevalence of AHD among clients who had CD4 count by sub-district**.

**S4 Fig. Overall number of PLWH who had scheduled visits and proportion of treatment interruption by district**.

**S5 Fig. Overall number of PLWH who had scheduled visits and proportion of treatment interruption by sub-district**.

**S6 Fig. Overall number of PLWH who had scheduled visits more than 5 months from ART initiation and proportion of viraemia by district**. VL, viral load; mL, millilitre.

**S7 Fig. Overall number of PLWH who had scheduled visits more than 5 months from ART initiation and proportion of viraemia by sub-district**. VL, viral load; mL, millilitre.

**S1 Table. Characteristics of clients by year and CD4 count category at ART initiation**.

**S2 Table. Characteristics of clients by scheduled visit year and treatment interruption status at first visit in each year**.

**S3 Table. Characteristics of clients by viral load year and viral load category at viral load testing**.

**S4 Table. Unadjusted relative risk of clinic-level prevalence of AHD, treatment interruption, viraemia**.

